# School-based caries prevention and the impact on acute and chronic student absenteeism

**DOI:** 10.1101/2022.11.22.22282638

**Authors:** Ryan Richard Ruff, Rami Habib, Tamarinda Barry-Godin, Richard Niederman

**Author notes:** Correspondence to: Ryan Richard Ruff, PhD, MPH, NYU College of Dentistry, 433 First Avenue, Room 712, New York, NY, 10010.

## Abstract

**Background:** Poor oral health is negatively associated with absenteeism, being attributed to millions of lost school days per year. The role of school-based dental programs that address oral health inequities on student attendance has not yet been explored.

**Methods:** Caried*Away* was a longitudinal, cluster-randomized, non-inferiority trial of preventive medicines for dental caries used in a school-based program. To explore the potential impact of caries prevention on attendance, we extracted data on average school absenteeism and the proportion of chronically absent students from publicly-available datasets maintained by the New York City Department of Education for years before, during, and after program onset. Data were obtained for all Caried*Away* schools as well as a group of untreated comparator schools. Total absences and the proportion of chronically absent students were modeled using multilevel mixed effects linear and two-limit tobit regression, respectively. Multiple model specifications were considered, including exposures to time-varying treatments across multiple years. Models also included a group of untreated comparator schools.

**Results:** In years in which treatment was provided through a school-based comprehensive caries prevention program, schools recorded approximately 944 fewer absences than in non-treatment years (95% CI = −1739, −149). Averaged across all study years, schools receiving either treatment had 1500 fewer absences than comparator schools, but this was not statistically significant. In contrast, chronic absenteeism was found to significantly decrease in later years of the program (B = -.037, 95% CI = -.062, -.011). Removing data for years affected by COVID-19 eliminated the significant reduction in total absences during treatment years, yet still showed a marginally significant interaction for chronic absenteeism.

**Discussion:** Though originally designed to mitigate access barriers to critical oral healthcare, early integration of school-based dental programs may positively impact school attendance. However, concerns over the reliability of attendance records due to the closing of school facilities resulting from COVID-19 may mask the true effect.

## Introduction

Dental caries is the most prevalent noncommunicative disease in the world and disproportionately affects children from disadvantaged backgrounds, such as those from low-income and/or minority families [1]. Children with poor oral health face numerous barriers to academic success including missed school days, lower test performance, difficulty paying attention, or impacts on functional and/or psychological behavior [2, 3]. In 2019, a systematic review and meta-analysis of studies in children aged 5-18 years concluded that poor oral health was associated with a significant increase in the risk of absenteeism and poor achievement [4]. In contrast, Medicaid children who receive comprehensive screening services early in life may demonstrate higher academic performance [5].

Despite the potential health and cognitive benefits of early childhood dental care, high-risk children often have lower dental service utilization rates [6-8]. Historical data from the US Department of Health and Human Services indicates that over 75% of children with Medicaid fail to receive required dental services, and one in four children do not see a dentist at all [9]. Partially to address this unmet need, multiple federal agencies including the Centers for Disease Control and Prevention recommend school-based dental programs, which can provide a range of effective preventive and/or therapeutic services and increase access to essential oral healthcare [10], improve oral health-related quality of life [11], and reduce the burden of disease [12, 13]. While utilization of general school-based health centers (e.g., medical, mental, dental, and vision services) may positively impact student attendance [14], the existing evidence is limited [15].

Caried*Away* is a pragmatic cluster-randomized trial of treatments for dental caries provided through a school-based program [16]. Conducted in predominantly low-income minority urban students, secondary objectives of the Caried*Away* trial were to assess the effects of school-based oral health programs on educational performance.

## Methods

### Design and Participants

Caried*Away* is a longitudinal, cluster-randomized, non-inferiority pragmatic trial of minimally-invasive interventions for dental caries implemented in schools. The study received ethical approval from the New York University School of Medicine Institutional Review Board (#i7-00578) and is registered at www.clinicaltrials.gov (#NCT03442309, 22/09/2018). A trial protocol was previously published [16]. Participant recruitment followed a two-stage process. First, any school with a student population consisting of at least 50% Hispanic/Latino or black race/ethnicity and at least 80% receiving free and reduced lunch was eligible for inclusion. Second, schools were randomized to treatments and any child in enrolled schools who provided parental informed consent, child assent, and spoke English was enrolled.

### Interventions

Schools were block-randomized using a random number generator to one of two experimental conditions: a “simple” experimental treatment consisting of fluoride varnish (5% NaF, Colgate PreviDent) applied to all teeth and a 38% silver diamine fluoride solution (Elevate Oral Care Advantage Arrest, 2.24 F-ion mg/dose) applied to any asymptomatic cavitated lesions and on all pits and fissures of bicuspids and molars, and a “complex” active comparator which included the same fluoride varnish application, glass ionomer sealants (GC Fuji IX) placed on all pits and fissures of bicuspids and molars, and atraumatic restorations on any frank asymptomatic cavitations.

### Comparator schools

For this analysis, a subset of schools which did not receive either simple or complex treatment assignment was included. This group consisted of schools that were enrolled in Stage 1 of the Caried*Away* recruitment process but did not proceed to stage two. These schools serve as non-randomized counterfactuals, in that they meet all study inclusion criteria, are found in the same geographic area, but have not received any treatment.

### Data Sources and Outcomes

Attendance rates for each included school were obtained for the 2016 through 2021 school years from the New York City Department of Education. Student attendance is attributed to the school the student attended at the time. If a student changed schools, attendance data was attributed to multiple schools. Data were extracted for total days present, total days absent, overall average school attendance, and the proportion of children classified as chronically absent. Chronic absenteeism was defined by the New York State Department of Education as any student with a total attendance of 90% or less across all school days (minimum enrollment threshold of ten days).

### Statistical Analysis

Data were ordered sequentially by school and year of study. Descriptive statistics for outcomes and select independent variables were produced overall and by treatment group. Absenteeism was modeled using mixed effects linear regression, and the proportion of chronically absent students using two-limit mixed effects tobit regression. Our first model (Model 1) was a single-group analysis defined as *y*_*t*_ = *b*_0_ + *b*_1_*t*_*t*_ + *b*_2_*x*_*t*_ + *b*_3_*x*_*t*_*t*_*t*_, where *t* is the overall time since the start of the Caried*Away* trial, *x* is a dummy variable indicating onset of the intervention (e.g., signifying a change from non-intervention period to an intervention period and vice-versa), and *xt* their interaction. Multiple treatment onsets were possible. Subsequent models introduced an additional parameter, *z*, representing the treatment type provided in each school (Model 2), and a series of treatment-specific interaction terms including the treatment-time interaction, treatment-intervention onset, and a three-level interaction between treatment, time, and intervention-onset, defined as *y*_*t*_ = *b*_0_ + *b*_1_*t*_*t*_ + *b*_2_*x*_*t*_ + *b*_3_*x*_*t*_*t*_*t*_ + *b*_4_*z* + *b*_5_*zt*_*t*_ + *b*_6_*zx*_*t*_ + *b*_7_*zx*_*t*_*t*_*t*_ (Model 3). Models were first run in schools receiving either simple or complex treatment (Models 1a, 2a, and 3a) and then inclusive of comparator schools, which did not receive treatment, by modifying the dummy indicator for treatment as ‘any treatment’ versus ‘no treatment’ (Models 1b, 2b, and 3b). Schools were included as random intercepts. As a final analysis, we excluded the 2019-2020 school year, which was partially conducted virtually due to the effects of the COVID-19 pandemic and may bias results.

## Results

When restricted to schools that received treatment in the Caried*Away* trial, our data included 193 yearly observations across 39 schools. When adding comparator schools, results reflect 52 schools and 258 yearly observations. The yearly recorded days absent and proportion of students chronically absent is shown in Table 1.

**Table 1:** Descriptive statistics for absenteeism and chronically absent students, by year

| Year | Simple |  |  |  | Complex |  |  |  | No treatment |  |  |  |
| --- | --- | --- | --- | --- | --- | --- | --- | --- | --- | --- | --- | --- |
|  | Days Missed |  | Chronically absent |  | Days Missed |  | Chronically absent |  | Days Missed |  | Chronically absent |  |
|  | Mean | SD | % | SD | Mean | SD | % | SD | Mean | SD | % | SD |
| 2016-2017 | 7600.53 | 3966.18 | 33.99 | 5.64 | 7200.33 | 3326.4 | 31.81 | 12.82 | 8221.85 | 3287.16 | 36.92 | 8.97 |
| 2017-2018 | 8360.41 | 4735.45 | 36.64 | 5.74 | 7373.76 | 3122.62 | 33.15 | 12.18 | 8275.77 | 3278.24 | 40.59 | 8.04 |
| 2018-2019 | 7988.44 | 4533.4 | 39.01 | 10.02 | 7276.1 | 3043.91 | 33.86 | 13.2 | 7995.85 | 2965.69 | 40.28 | 7.73 |
| 2019-2020 | 4718.72 | 2300.48 | 34.03 | 7.42 | 4352.48 | 1767.06 | 31.81 | 11.38 | 4606.54 | 1773.5 | 35.52 | 9.22 |
| 2020-2021 | 7660.28 | 3677.6 | 36.71 | 12.53 | 7933.95 | 3859.62 | 37.61 | 14.74 | 8089.39 | 3616.8 | 39.1 | 15.13 |

Results for total days absent (Table 2) and chronic absenteeism (Table 3) combine models for simple versus complex schools (Model 1a, Model 2a, Model 3a) and any treatment versus no treatment schools (Model 1b, Model 2b, Model 3b). For total days absent, there was a consistent reduction in school absence in years in which treatment was provided (‘x’ indicator) as well as an increased effect when treatment was provided in later school years (‘xt’ indicator). For example, Model 2b with all schools included shows a predicted reduction of 883 (95% CI = −1697, −70) missed school days in years in which treatment was provided with an additional reduction of 1437 days (95% CI = −2219, −655) if treatment was provided in later years. Including all schools and relevant predictors (Model 3b), there was an additional non-significant overall reduction in missed school days in schools that ever received treatment versus schools that did not (B=-1505, 95% CI = −3699, 690), but this gap significantly reduced over time (B=556, 95% CI = 221-890). Indicators for the treatment/treatment time and treatment/treatment time/overall time variables were perfectly collinear as comparator schools never received treatment and were therefor removed from the final model.

**Table 2:** Model results for total school absences

| VARIABLES | (1)<br>Model 1a | (1)<br>Model 1b | (2)<br>Model 2a | (2)<br>Model 2b | (3)<br>Model 3 | (3)<br>Model 3b |
| --- | --- | --- | --- | --- | --- | --- |
| t | 137.8<br>(-33.08 - 308.8) | -25.20<br>(-180.8 - 130.4) | 138.0<br>(-32.91 - 308.9) | -24.75<br>(-180.5 - 131.0) | 63.23<br>(-198.3 - 324.8) | -418.3***<br>(-699.7 - -136.9) |
| x | -945.2**<br>(-1,698 - -192.0) | -879.9**<br>(-1,692 - -67.34) | -946.0**<br>(-1,699 - -192.8) | -883.5**<br>(-1,697 - -69.68) | -1,037**<br>(-2,064 - -10.51) | -943.9**<br>(-1,739 - -149.1) |
| xt | -1,607***<br>(-2,337 - -877.9) | -1,437***<br>(-2,219 - -654.7) | -1,609***<br>(-2,339 - -879.7) | -1,437***<br>(-2,219 - -655.1) | -1,135**<br>(-2,060 - -210.1) | -1,608***<br>(-2,378 - -837.9) |
| z |  |  | -534.2<br>(-2,554 - 1,486) | 154.7<br>(-1,802 - 2,111) | -745.2<br>(-2,984 - 1,493) | -1,505<br>(-3,699 - 689.6) |
| zt |  |  |  |  | 125.4<br>(-217.4 - 468.1) | 555.6***<br>(221.4 - 889.9) |
| zx |  |  |  |  | 4,129<br>(-1,535 - 9,792) |  |
| zxt |  |  |  |  | -1,232<br>(-2,724 - 259.0) |  |
| Constant | 7,238***<br>(6,120 - 8,355) | 7,632***<br>(6,676 - 8,587) | 7,526***<br>(5,968 - 9,084) | 7,515***<br>(5,759 - 9,271) | 7,650***<br>(5,998 - 9,302) | 8,744***<br>(6,844 - 10,645) |
| Observations | 193 | 258 | 193 | 258 | 193 | 258 |
| Schools | 39 | 52 | 39 | 52 | 39 | 52 |

**Table 3:** Model results for the proportion of chronically absent students

| VARIABLES | (1)<br>Model 1a | (1)<br>Model 1b | (2)<br>Model 2 | (2)<br>Model 2b | (3)<br>Model 3 | (3)<br>Model 3b |
| --- | --- | --- | --- | --- | --- | --- |
| t | 0.0114***<br>(0.00542 - 0.0174) | 0.00766***<br>(0.00254 - 0.0128) | 0.0114***<br>(0.00543 - 0.0174) | 0.00757***<br>(0.00245 - 0.0127) | 0.00625<br>(-0.00296 - 0.0155) | -0.00184<br>(-0.0112 - 0.00751) |
| x | 0.00200<br>(-0.0244 - 0.0284) | 0.00268<br>(-0.0240 - 0.0294) | 0.00194<br>(-0.0245 - 0.0283) | 0.00343<br>(-0.0233 - 0.0302) | 0.00846<br>(-0.0277 - 0.0446) | 0.00200<br>(-0.0244 - 0.0284) |
| xt | -0.0369***<br>(-0.0624 - -0.0113) | -0.0329**<br>(-0.0586 - -0.00723) | -0.0370***<br>(-0.0626 - -0.0114) | -0.0328**<br>(-0.0585 - -0.00715) | -0.0328**<br>(-0.0654 - -0.00026) | -0.0369***<br>(-0.0624 - -0.0113) |
| z |  |  | -0.0283<br>(-0.0900 - 0.0334) | -0.0285<br>(-0.0890 - 0.0319) | -0.0498<br>(-0.120 - 0.0205) | -0.0681*<br>(-0.137 - 0.00108) |
| zt |  |  |  |  | 0.00885<br>(-0.00322 - 0.0209) | 0.0133**<br>(0.00216 - 0.0244) |
| zx |  |  |  |  | 0.00758<br>(-0.192 - 0.207) |  |
| zxt |  |  |  |  | -0.00643<br>(-0.0589 - 0.0461) |  |
| Constant | 0.322***<br>(0.287 - 0.358) | 0.340***<br>(0.310 - 0.370) | 0.338***<br>(0.289 - 0.386) | 0.361***<br>(0.307 - 0.416) | 0.350***<br>(0.298 - 0.402) | 0.391***<br>(0.331 - 0.451) |
| Observations | 193 | 258 | 193 | 258 | 193 | 258 |
| Schools | 39 | 52 | 39 | 52 | 39 | 52 |

For the proportion of enrolled schoolchildren who are chronically absent, single-group models (Table 3, Model 1a and 1b) indicate that there is a significant yearly increase of approximately 1% in the proportion of chronically absent students each year, and a significant 3% decrease for the interaction between school year and if the school received treatment that year. Multi-group models with and without comparator schools show a non-significant reduction in chronic absenteeism for simple versus complex treatment (Model 2a) and any treatment versus no treatment (Model 2b). Overall model results (Model 3b, inclusive of comparator schools) suggests that there is an approximate 3.5% decrease in chronic absenteeism when treatment was provided in later years. There was a marginally significant (p=.054) decrease of 7% in schools receiving treatment compared to control schools, but this effect reduces by 1% over time.

Results when excluding data from the 2019-2020 school year removed the effect of treatment in a given year. There was an overall reduction in days absent comparing treated schools to untreated schools, but this effect was not statistically significant (Table 4, Models 1 & 2. Similar results were found for chronically absent students, with a non-significant reduction in the chronically absent population in treated schools (Table 4, Models 3 & 4).

**Table 4:** Model results for absences (Models 1 and 2) and chronically absent students (Models 3 and 4), excluding remote learning years

| VARIABLES | (1)<br>Model 1 | (2)<br>Model 2 | (3)<br>Model 3 | (4)<br>Model 4 |
| --- | --- | --- | --- | --- |
| t | 58.01<br>(-56.19 - 172.2) | 68.75<br>(-48.15 - 185.7) | 0.00795***<br>(0.00244 - 0.0135) | 0.00912***<br>(0.00353 - 0.0147) |
| x | 32.45<br>(-597.1 - 662.0) | 127.3<br>(-541.6 - 796.3) | -0.00631<br>(-0.0365 - 0.0238) | 0.00431<br>(-0.0275 - 0.0362) |
| z | -550.7<br>(-2,703 - 1,601) | -545.0<br>(-2,695 - 1,605) | -0.0363<br>(-0.0983 - 0.0256) | -0.0357<br>(-0.0975 - 0.0261) |
| xt |  | -277.8<br>(-949.1 - 393.5) |  | -0.0305*<br>(-0.0625 - 0.00142) |
| Constant | 7,977***<br>(6,086 - 9,868) | 7,946***<br>(6,055 - 9,837) | 0.369***<br>(0.313 - 0.425) | 0.366***<br>(0.310 - 0.422) |
| Observations | 206 | 206 | 206 | 206 |
| Schools | 52 | 52 | 52 | 52 |

ci in parentheses \*\*\* p<0.01, \*\* p<0.05, \* p<0.1

## Discussion

In addition to school absences resulting from dental pain or infection, acute or unplanned dental care contributes over thirty million hours of missed school per year [17]. Although school-based dental programs were developed to treat and prevent oral diseases in children who lack access to traditional dental care [18], their early integration may likewise improve educational performance. The Caried*Away* project provided both comprehensive dental screening/examinations and treatment services for untreated dental caries, supporting preliminary analyses of early school-based dental interventions on academic outcomes.

Prior research suggests that the utilization of school-based health centers positively affects classroom attendance and seat time, reflecting the complex role of child health on education [19, 20]. Our findings indicate that school-based dental care may reduce student absenteeism, but the changes in school operations due to COVID-19 could bias results. Inclusive of all years, there was an average reduction of 940 missed school days in years that treatment was provided. There were also no differences in absenteeism when comparing schools receiving simple or complex prevention. For this latter finding, prior data from Caried*Away* indicated that the clinical effectiveness of simple prevention was non-inferior to that of complex prevention, with near identical rates of disease prevention over time [12]. As both treatments in the Caried*Away* project had similar impact, non-differential effects on secondary outcomes, such as absenteeism, may be expected.

Many schools participating in the Caried*Away* program received treatment during the 2019-2020 school year; as a result, the effect of treatment in a given year may be biased due to the impact of COVID-19. Schools in New York City closed educational facilities and transitioned to remote learning on 15 March 2020, and the final third of the school year was conducted virtually. On the one hand, attendance and achievement may decrease when transitioning to remote instruction, with low-income areas being particularly affected [21]. Indeed, publicly-accessible data from 1,500 schools in New York City indicated that schools with heavy Black and Hispanic populations were much more likely to report poor attendance during remote instruction [22]. On the other, the administrative confusion associated with such an unprecedented transition to remote learning may have resulted in inaccurate or unreliable reporting, and there was a noticeable drop city-wide in absenteeism during this initial period.

We first attempted to explore this potential bias by including schools enrolled in Caried*Away*, but not treated, in our analysis. Compared to these schools, treated schools did record an additional reduction of approximately 1,500 missed school days, but this effect was not statistically significant. However, this indicator reflects average attendance throughout all years of the program, inclusive of years care was not provided in treatment schools. Additionally, some schools received care in multiple years, including years not affected by remote learning policies. We then further restricted our data to excluded attendance records for the 2019-2020 school year. In so doing, we found that the previously estimated reduction in absences was removed. However, there remained a consistent, non-significant reduction in overall absences and when comparing treated to untreated schools.

In contrast to accumulated absences, data reporting for the proportion of students who were chronically absent may be more robust to the effects of remote instruction, particularly as this transition occurred in the last quarter of the academic year. While treatment onset did not have an immediate impact on this outcome (e.g., reductions in the same year in which treatment was provided), there was a significant interaction for subsequent years and treated schools showed a marginally significant reduction compared to untreated ones. When removing the 2019-2020 school year, we similarly found a non-significant reduction in the chronically absent population in treated schools. Recently, the 2022 Mayor’s Management Report documented a rise in city-wide chronic absenteeism, citing continued disruptions due to COVID-19 variants and a re-transition to in-person instruction [23]. As Caried*Away* prioritized schools with predominantly low-income, minority student populations, our findings could suggest a potential positive pathway to improved attendance via those students who are chronically absent.

In spite of concerns regarding the effects of COVID-19 and the shifts in educational instruction, our analysis suggest that school-based comprehensive dental programs could potentially have a positive effect on attendance and chronic absenteeism. Indeed, the overall participation rate across all schools enrolled and treated in Caried*Away* was approximately 30%. As a result, the true impact on school-level absenteeism may be further masked by children who elected not to participate. Further study using student-level data may show the potential pathways to affecting attendance, such as analysis stratified by baseline decay in children or exploring the post-treatment longitudinal effects, such as attendance trajectories for children from early grades through primary school graduation.

## Data Availability

Data for the present work are available upon reasonable request to the authors.

## Author Contributions

RRR and RN conceived of the study, obtained funding, and were the Principal Investigators of the Caried*Away* trial. RRR performed all statistical analyses and wrote the manuscript. RH collected educational data. TBG directed the Caried*Away* program and collected clinical data. All authors critically reviewed and revised the manuscript.

